# Preliminary Results of Seroprevalence of SARS-CoV-2 at Community Clinics in Tokyo

**DOI:** 10.1101/2020.04.29.20085449

**Authors:** Morihito Takita, Tomoko Matsumura, Kana Yamamoto, Erika Yamashita, Kazutaka Hosoda, Tamae Hamaki, Eiji Kusumi

**Author notes:** Corresponding Author: Morihito Takita, Department of Internal Medicine, Navitas Clinic Tachikawa, Address: 3-1-1 Shibasaki, Ecute Tachikawa 4^th^ floor, Tachikawa, Tokyo, 190-0023, Japan. Conflict of Interest We declare no competing interests. The study was performed by intramural funding of Navitas Clinic.

## Abstract

Serological evaluation with SARS-CoV-2 specific IgG antibody will be an alternative way to know the pandemic of novel coronavirus disease (COVID-19) if the capacity for diagnostic PCR test is limited. The point-of-care test to detect SARS-CoV-2 specific IgG antibody in peripheral blood (*n* =202) was performed in two community clinics in Tokyo, Japan. The overall positive rate of SRAS-CoV-2 IgG antibody was 5.9% (95% confidence interval[CI]: 3.1-10.1). Higher rate was observed for healthcare workers (*n* =55, 9.1 [3.0-20.0]). The limitation on antibody tests includes low sensitivity and potent cross-reactivity with the previous coronavirus. Robust healthcare policy to efficiently monitor COVID-19 spread is warranted in Tokyo.

## Main text

Delay in the expansion of the capacity for diagnostic PCR test to detect severe acute respiratory syndrome coronavirus 2 (SARS-CoV-2) in Japan has made difficulties in evaluating the pandemic of novel coronavirus disease (COVID-19), in turn, planning appropriate public measures.^1^ Serological evaluation with SARS-CoV-2 specific IgG antibody will be an alternative way to know how many people have been infected in the individual region.^2^ The State of New York recently released the preliminary results of antibody tests showing that the SARS-CoV-2 specific IgG antibody was detected in approximately 15% of the citizens.^3^ Of note, the WHO made a caution on the concept of Immunity Passport.^4^

We here report our preliminary results of the SARS-CoV-2 specific IgG antibody measured by the point-of-care immunodiagnostic test (SARS-CoV-2 Antibody Testing Kit IgG RF-NC002, Kurabo Industries Ltd, Osaka, Japan) in the two community clinics located in the major railway stations in Tokyo (Navitas Clinic Shinjuku at Shinjuku Station and Navitas Clinic Tachikawa at Tachikawa Sta.). The Institutional Review Board of Navitas Clinic approved the study (Approval Number: NC2020–01). Asymptomatic subjects have been recruited by web posting of our clinic, and written consent was obtained prior to the test.

A total of 202 participants, including 55 healthcare workers (physicians, nurses, pharmacists, and laboratory technicians), participated in the study between April 21 and 28, 2020 (Table).

Fifty-two participants (26%) experienced an episode of fever within a month of antibody test, whereas the PCR test was performed for 9 (4%). The positive rate of SRAS-CoV-2 IgG antibody was 5.9% (95% confidence interval[CI]: 3.1-10.1), consisting of six males (4.9% [1.8-10.3]) and six females (7.6% [2.8-15.8]). A healthcare worker who was only a participant with a positive result of the PCR test showed positive for IgG. The regional difference of the antibody-positive rate is 6.7% (95% CI: 3.4-11.6) and 2.7% (0.1-14.2) in Shinjuku of central Tokyo and Tachikawa of the suburban, respectively.

A significant issue of pandemic COVID-19 is how we can identify and manage asymptomatically infected people, who may cause a chain of epidemic contagion and nosocomial infection in medical institutions, along with intensive medical care for severely symptomatic patients.^5^ The limitation on antibody tests includes low sensitivity and potent cross-reactivity with the previous coronavirus. Our results, however, suggest a higher population infected with COVID-19 in metropolitan Tokyo than the national survey daily updated.^6^ Robust healthcare policy to efficiently monitor COVID-19 spread is warranted in Tokyo.

Table

|  | Healthcare Workers<br>(n =55) | Non-Healthcare<br>Workers (n =147) | Total (n =202) |
| --- | --- | --- | --- |
| Age(year) | n (%) | n (%) | n (%) |
| 20-29 | 0 (0) | 9 (6) | 9 (4) |
| 30-39 | 5 (9) | 30 (21) | 35 (17) |
| 40-49 | 20 (36) | 58 (39) | 78 (39) |
| 50-59 | 9 (16) | 30 (21) | 39 (19) |
| 60-69 | 17 (31) | 15 (10) | 32 (16) |
| 70-80 | 4 (7) | 5 (3) | 9 (4) |
| Gender-Female | 19 (35) | 60 (41) | 79 (39) |
| History of fever within a month | 10 (18) | 42 (29) | 52 (26) |
| Existence of cohabitant diagnosed<br>with COVID-19 | 0 (0) | 2 (1) | 2 (1) |
| History of PCR test for COVID-19 | 2 (4) | 7 (5) | 9 (4) |
| Positive Rate of SARS-CoV-2 IgG<br>antibody -n, (% , [95% Confidence<br>Interval]) | 5 (9.1 [3.0-20.0]) | 7 (4.8 [1.9-9.6]) | 12 (5.9 [3.1-10.1]) |

## Data Availability

The datasets analyzed during the current study are not publicly available due to ongoing further analyses but are available from the corresponding
author on reasonable request.

